# SleePyPhases: A Workflow Framework for Sleep Data Harmonization, Analysis and Machine Learning

**DOI:** 10.64898/2026.01.14.26344163

**Authors:** Franz Ehrlich, Sara Bäcker, Martin Schmidt, Hagen Malberg, Martin Sedlmayr, Miriam Goldammer

## Abstract

Data-driven sleep research relies on polysomnography data from various public repositories and vendor systems. Yet the lack of standardized access methods creates substantial barriers to multi-dataset research, method reuse, and reproducibility. We present SleePyPhases, an open-source Python framework providing unified access to multiple sleep data repositories. It offers integrated data harmonization, configuration-based preprocessing, and the development of machine learning pipelines. The framework unifies channel naming, annotation semantics, and data formats across several public repositories (including SHHS, MESA, MrOS, PhysioNet, and SleepEDF) and commercial vendor formats (Philips Alice and Somnomedics Domino). We validated the framework by reproducing five published sleep analysis studies covering diverse datasets, sleep scoring tasks (sleep staging, arousal, leg movement, respiratory event detection), preprocessing methods (signal preprocessing and spectrograms), machine learning methods (supervised and unsupervised learning), and model architectures (convolutional, recurrent, and transformer networks). Four reproductions achieved near-identical results, confirming data fidelity and methodological flexibility. SleePyPhases is open-source and provides a foundation for reproducible sleep research, enabling researchers to focus on scientific questions rather than data infrastructure.

## 1. Introduction

Sleep is a fundamental aspect of human health and well-being (Mukherjee et al. 2015) and plays a crucial role in various physiological and cognitive processes. Understanding the complexities of sleep has therefore been the focus of numerous research studies, providing valuable insights into the prevention, diagnosis and treatment of sleep disorders. Polysomnography (PSG), performed in accredited sleep laboratories, is the gold standard for assessing sleep quality. These overnight recordings contain at least nine different types of biosignals, along with manual annotations and metadata (Berry et al. 2012). Biosignals are measured using multiple derivations and are often supplemented by additional signals, such as those from positive airway pressure therapy devices. Sleep technologists and physicians manually score the full-night PSG, adding annotations for sleep stages, arousals, respiratory events and leg movements. Sleep laboratories can choose from numerous software suppliers to record and store PSG data.

However, the lack of standardization in the sleep research field poses significant challenges for researchers and clinicians alike (Mazzotti et al. 2022; Zhang et al. 2018).

Due to this absence of standards, manufacturers use proprietary formats for storing sleep data, making it difficult to exchange, analyze, and process data from different sources. Although the European Data Format (EDF+) (Kemp and Olivan 2003) provides a standardized file format for storing biosignals, it lacks semantic meaning in terms of channel names and annotation labels, which are crucial for consistent interpretation and analysis across studies. These factors make it difficult to create automated data analysis pipelines that work across different datasets, vendors, or PSG configurations.

To gain a comprehensive understanding of sleep, researchers rely on the analysis of large cohorts containing sleep-related information, such as polysomnography (PSG) recordings (Quan et al. 1997; Chen et al. 2015; Blackwell et al. 2011). These datasets, often collected from diverse populations and settings, have the potential to reveal patterns, associations and trends that can significantly advance our knowledge of sleep. Machine learning and “big data” techniques, which have shown great promise in sleep research (Redline and Purcell 2021), require consistent input data, with all channels having the same sampling rate across recordings. This harmonisation process is essential for developing robust and generalisable models that can leverage the full potential of large-scale sleep datasets.

The lack of standardization in sleep research has been widely recognized as a significant barrier to cross-study analysis and reproducibility (Zhang et al. 2018; Arnardottir et al. 2016). There exist only few publications that address these problems and are therefore a relevant comparison to this work.

Zhang et al. highlighted the critical need for data sharing and reuse of data to accelerate sleep research. Therefore they created a space to store digital objects and made them **Findable, Accessible, Interoperable and Reusable** (FAIR principle (Wilkinson et al. 2016)). The space includes multiple sleep datasets emphasising the EDF format and standard XML files for annotations and CSV for metadata. Their main goal of Zhang et al. (2018) was to create a comprehensive set of harmonized data. However, this does not include a harmonized method for loading and processing the data.

Sveinbjarnarson et al. (Sveinbjarnarson et al. 2023) described the “Sleep Revolution Platform,” proposing a high-level design for dynamic data source pipelines and digital platform architecture for complex sleep data. Their work emphasizes the theoretical framework for transforming heterogeneous datasets into homogeneous databases and discusses database management considerations. However, unlike the present work, their contribution remains primarily at the design level without a complete implementation or extensive validation across diverse machine learning approaches.

Recent efforts to address the reproducibility of sleep-related research include SLEEPYLAND (Fiorillo et al. 2024), which provides a comprehensive framework for sleep data analysis, emphasising the fair training of sleep staging models locally. The project offers a graphical user interface (GUI) for selecting various state-of-the-art algorithms. These models can be executed locally using multiple publicly available datasets or local EDF data. The GUI also offers a range of visualisation methods for evaluating the algorithms. The aim of SLEEPYLAND is to fairly retrain existing methods.

One recent study has specifically addressed the challenges of multi-dataset sleep research by using standardized data pipelines. Strøm et al. (Strøm et al. 2024) developed the “Common Sleep Data Pipeline” (CSDP), an open-source solution for standardizing heterogeneous sleep datasets into a unified format using HDF5 storage.

Their approach demonstrates processing 21 datasets, with validation through U-Sleep and L-SeqSleepNet models. While the CSDP primarily focuses on sleep staging, data harmonisation, and storage efficiency, it lacks other sleep scoring tasks, extensive preprocessing, machine learning workflow integration, and customizable vendor format support.

There are many toolboxes and packages for sleep analysis in Python, including a U-Sleep implementation that supports multiple datasets (Perslev et al. 2021). However, as these prioritise reproducible analysis steps over an overarching framework, they are not relevant to this work.

With a focus on enabling reproducible, multi-dataset machine learning research, we have developed the workflow framework *SleePyPhases* in Python. This modular, extensible framework provides:

- **Unified data access** from proprietary formats and publicly available datasets through standardized interfaces
- **Semantic harmonization** of channel names, annotation labels, and metadata across diverse sources
- **Configuration-driven workflows** that ensure reproducibility and minimal data generation
- **Extensible plugin architecture** enabling community contributions for new data sources
- **FAIR-compliant** supporting findability, accessibility, interoperability, and reusability

To validate that the workflow framework, we reproduced five published sleep analysis from various domains of sleep analysis and reproduced their results. The reproductions span a wide range of:

- Publicly available datasets including SHHS, MESA, MrOS, PhysioNet and SleepEDF
- Single and multi-task sleep analysis including sleep staging, arousal detection, sleep disordered breathing detection and leg movement detection
- Different labeling strategies such as default event windows and segmentation
- Different preprocessing requirements such as signal-based and spectrogram-based approaches
- Different model architectures including CNN, LSTM and Transformer
- Different training strategies such as supervised and unsupervised learning

The aim of *SleePyPhases* is to provide a standardised workflow for accessing sleep data that is independent of repository, vendor or study design. The validation cases presented demonstrate the flexibility of the applied methods while reducing the technical overheads of multi-dataset research.

## 2. Materials and Methods

The *SleePyPhases* workflow framework was implemented in Python to leverage the ecosystem of scientific computing tools. The framework evolved through iterative application to our research projects (Goldammer et al. 2022; Ehrlich et al. 2022, 2024). The modular architecture emerged as a necessity to accommodate the inherent heterogeneity of sleep data across repositories, vendors, and research methodologies.

This section describes the framework’s design principles, harmonization approach, and validation through reproduction of five diverse published studies.

### 2.1 Framework Requirements and Design Objectives

The development of machine learning pipelines for sleep data analysis presents several fundamental challenges that must be addressed to enable reproducible and scalable research. Based on the literature (Cheng et al. 2024), our previous development process and experience with diverse sleep analysis approaches, we identified the following core requirements for a comprehensive sleep analysis framework:

#### Data Harmonization

The framework must provide unified access to a variety of sleep data sources, including both proprietary vendor formats and publicly available datasets. This requires reconciling heterogeneity across three dimensions: syntax (technical formats like XML or JSON), structure (conceptual schemas, such as transforming signal-based annotations into event-based data), and semantics (the intended meaning of variables) (Cheng et al. 2024). This process is centered on the definition of a DataSchema (Cheng et al. 2024). For sleep research this means channel naming conventions, annotation semantics and PSG parameters. Additionally, for machine learning tasks, a consistent temporal resolution is required.

#### Configurable Preprocessing

Preprocessing pipelines must be fully configurable, with efficient storage mechanisms that eliminate redundant computations. The system should support signal-type-specific preprocessing steps. This aligns with the requirement for normalization and standardization of data methodologies to ensure that different datasets are inferentially equivalent before being pooled for machine learning (Cheng et al. 2024).

#### Flexible Data Manipulation

Different machine learning algorithms require different input representations. The framework must provide extensible mechanisms for transforming preprocessed data for each training step.

#### Integrated Training Pipeline

The framework should integrate data loading, model training, and evaluation within a unified environment. This includes support for multiple deep learning frameworks, custom training strategies, and comprehensive evaluation metrics for both segment-wise and event-wise analysis.

#### Configuration-Data Synchronization

Every methodological decision must be transparently documented (Cheng et al. 2024). Method decisions should be documented using the configuration. To prevent inconsistencies and redundant computations, the framework should maintain explicit connections between configurations and generated artifacts. When configuration parameters change, the system should automatically recognize which artifacts need regeneration.

#### Modular Architecture Through Plugins

To address the logistical challenge of disparate stakeholders (different vendors, datasets, and research protocols) and evolving technology (Cheng et al. 2024) the framework must employ a modular plugin-based architecture. This enables parallel development of dataset-specific components and research methods while preserving reproducibility through versioned plugins that can be independently developed, tested, and shared across research groups.

To address these requirements, we developed *SleePyPhases* as a modular, extensible Python framework built upon three main components: the core pyPhases system for configuration-driven project management, the SleepHarmonizer plugin for PSG data handling, and the pyPhasesML plugin for machine learning operations. This architecture enables researchers to focus on algorithmic innovation rather than infrastructure concerns while ensuring that complex pipelines remain reproducible. By design, the framework adheres to FAIR principles (Wilkinson et al. 2016): it is **Findable** through public repositories, **Accessible** via open-source licensing, **Interoperable** across diverse PSG vendors, datasets and formats, and **Reusable** through its modular, configuration-driven architecture.

### 2.2 Framework Validation Through Study Reproduction

To validate that the harmonized data access workflow provides functionally equivalent data to direct repository access, we selected five published sleep analysis studies for reproduction. These studies were specifically chosen to comprehensively test the framework’s capability to handle diverse data access requirements and methodological approaches. Successful reproduction with near-identical results serves as validation that: (1) the harmonization workflow correctly loads and processes data from different repositories, (2) the standardized access layer does not introduce artifacts or degrade data quality, (3) the configuration-driven approach maintains reproducibility, and (4) previously closed-source methods can be made open and reproducible. Each reproduction required implementing data access through our framework rather than original custom code, directly testing whether standardized access can replace repository-specific implementations. An overview of the validation studies is given in Table 1.

**Table 1:**
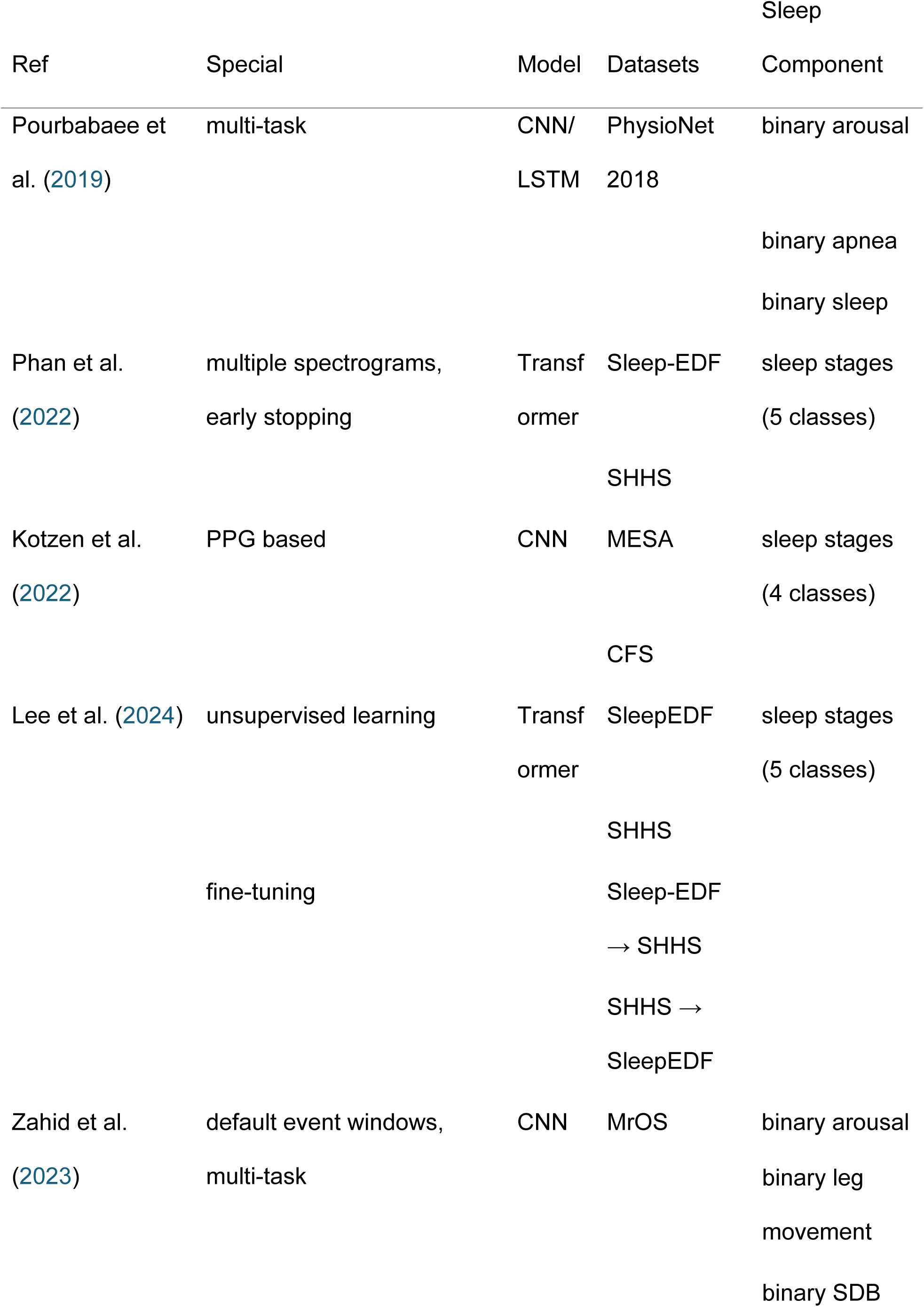
An overview of the selected research papers for framework validation. The papers cover a wide range of methodologies and datasets. SDB: Sleep Disordered Breathing CFS: Cleveland Family Study Visit-5 v1, SHHS: Sleep Heart Health Study, MESA: Multi-Ethnic Study of Atherosclerosis, MrOS: MrOS Sleep Study

A dense recurrent convolutional neural network (DRCNN) is used in the approach by (Howe-Patterson, Pourbabaee, and Benard 2018), which won the George B. Moody PhysioNet Challenges 2018 “You Snooze You Win” (Ghassemi et al. 2018) for detecting non-apnea arousals. Their approach includes a multi-task learning approach that combines binary classification for arousal, sleep and apnea events. The binary output is then remapped to only relevant states. The training pipeline, including the trained models, is available as open source.

The *SleepTransformer* implementation from (Phan et al. 2022) aims to create a more interpretable five-class sleep stage classification using a transformer-based architecture. Phan et al. calculate spectrogram images for the EEG, EOG and EMG in the SleepEDF and SHHS dataset. To reach better results on the much smaller SleepEDF dataset a pre-trained model trained on the SHHS data is used to improve the results. The open-source repository includes MATLAB code for preprocessing and evaluation purposes. Meanwhile, the training code is provided in Python.

*SleepPPG-Net* was introduced by Kotzen et al. (2022), a deep learning algorithm for four-class sleep staging directly from the continuous raw PPG time series. It utilizes a residual convolutional network for feature extraction and a temporal convolutional network for contextual information. It was developed and evaluated using the SHHS, MESA, and CFS public sleep databases. The different datasets are used for pretrained models using the electrocardiogram (ECG), inter-dataset validation and fine-tuning the model on a specific dataset.

The approach from (Zahid et al. 2023) proposes a CNN/RNN based split-stream architecture designed for multi sleep event detection (*MSED*) including arousals, leg movements, and binary sleep-disordered breathing events. The model was trained and tested on the MrOS Sleep Study, using various PSG channels. Compared to other approaches, default event windows are used. Instead of using fixed segmentation, they generate default events with different window sizes (3, 15, 30 seconds) across the whole record as target. The training task involves predicting the correct class and location for each default event window. To handle the class imbalance favoring non-class default events, negative mining loss was employed. A custom sampling strategy was used to select segments based on the event probability distribution across a recording to prevent segments without events being used. For the evaluation only events with at least an intersection over union (IOU) of 0.5 are compared.

*NeuroNet* (Lee et al. 2024) introduces a novel self-supervised learning (SSL) framework combining contrastive learning and masked prediction tasks with a Mamba-based temporal context module, all using single-channel EEG. The research uses three different datasets: SleepEDFX, SHHS, and ISRUC-Sleep (Khalighi et al. 2016). First an encoder with a representation of an EEG segment was learned using SSL. A total of five data augmentation methods were used, two of which were randomly selected during training. The training progress was monitored on a simple sleep staging task using a principal component analysis on the learned features during the validation. Next a simple classifier that predicts sleep stages on a single epoch was trained using the frozen encoder to evaluate the SSL performance. Afterwards a more comprehensive model was trained, that also included a temporal context of multiple sleep epochs. The training pipeline is available as open source, without the loading of the data and data augmentation.

### 2.3 Framework Implementation

#### 2.3.1 Core Architecture: pyPhases

The core of the SleePyPhases framework is pyPhases, a framework for creating and executing custom *Projects*. Its key feature is synchronizing generated data (e.g. preprocessed data and trained models) with the configuration. A *Project* is defined by three components:

*Phases*: These are used to execute code and generate data. Each phase can be executed separately. For a machine learning project, typical phases include data preprocessing, model training and evaluation.

*Configuration*: The Configuration defines the execution. Typical configuration for a machine learning task includes a target sampling rate, preprocessing steps, model hyperparameters and training parameters.

*Data*: Data is generated by the *Phases* and may depend on the *Configuration*. Data that depends on the configuration is stored with a specific configuration identification. For example preprocessed data is generated by the preprocessing phase and depends on the target sampling rate defined in the configuration.

These three components are defined in a project.yaml file, which provides a human readable project configuration. A basic example of project configuration and python code can be found in the appendix.

Another integral component of pyPhases is data storage. The storage is handled by an *Exporter*, which can store and load data. The most basic type of storage is a pickle-based *Exporter*, which handles the loading and storing of data directly to the filesystem.

With this construct of pyPhases, it is possible to generate data based on the *Project* state: When data depends on the configuration, it is stored with a specific configuration identification. When data was previously generated with the same configuration it does not need to be regenerated and can be directly loaded from the storage. Figure 1 shows an example of how a *Project* instance handles a data request.

**Figure 1:** An example of how the project instance handles a data request. The project checks if the data exists in memory or file-system and if not, it generates the data using the defined phases and saves the data using the defined exporter and stores it in memory. The data can be loaded from memory or file-system if it exists for the specific configuration.

Each *Project* comprises its own *Phases*, *Configuration*, *Data* and *Exporter*. The *pyPhases* framework also provides a plugin system, which helps to compose and extend different functionality. Each plugin is represented by a Python package in the *Python Package Index* and can be developed separately.

#### 2.3.2 SleePyPhases Architecture

The *SleePyPhases* architecture is specifically designed to create pipelines for automatic sleep analysis with machine learning. The overall architecture is shown in Figure 2. The complete training and evaluation workflow of a ML algorithm is shown in Figure 3, which illustrates how the *Phases* and modules interact to generate temporary and persistent data.

**Figure 2:**
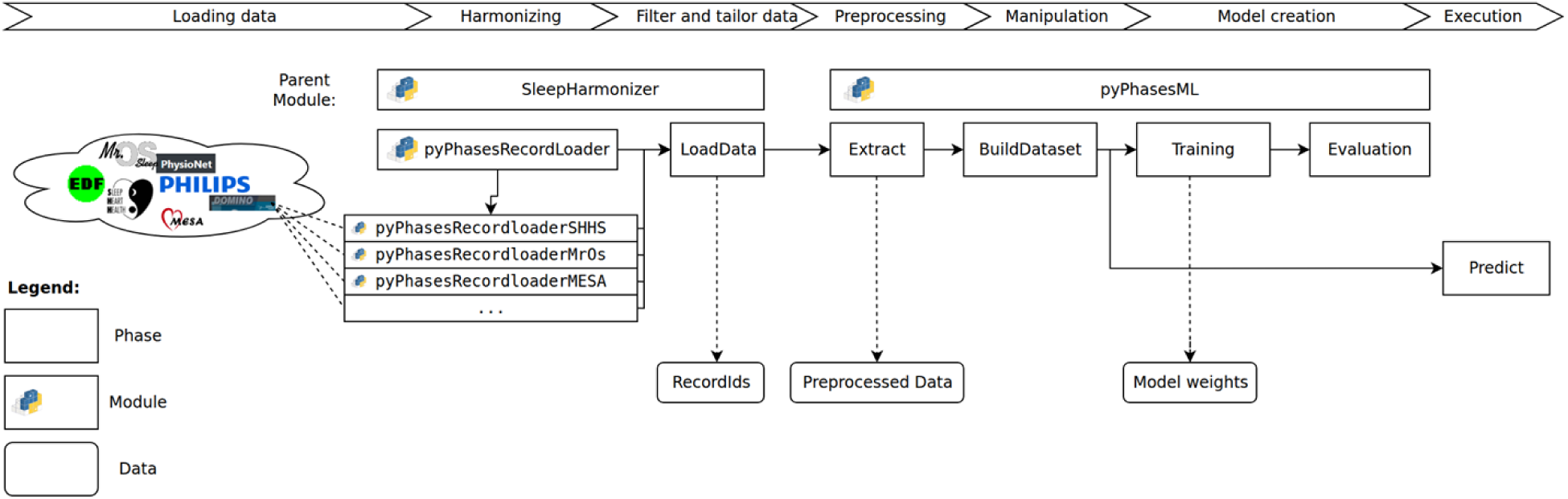
SleePyPhases Architecture

**Figure 3:**
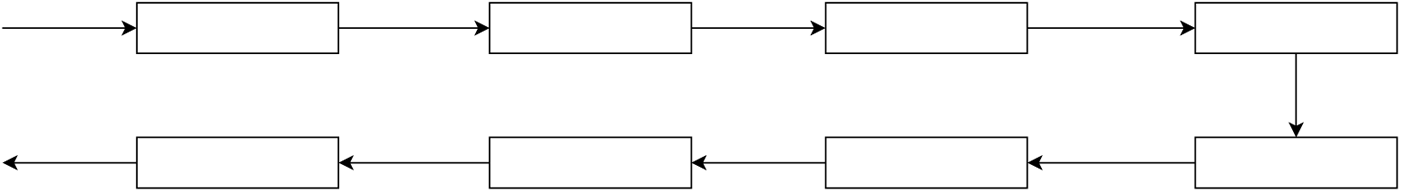
A sequential order for a machine learning model evaluation, where the Phases and modules produce different types of temporary and persistent (shown in bold) data.

The base functionality of *pyPhases* is extended by the plugins *pyPhasesML* and *SleepHarmonizer*. The *SleepHarmonizer* provides a standard interface for working with recordings using the pyPhasesRecordloader module. This module forms the basis for specific record loaders that support particular file types, datasets (e.g. pyPhasesRecordloaderSHHS) or PSG vendors (e.g. pyPhasesRecordloaderDomino). These specific record loaders provide an interface that harmonizes different biosignals, manual annotations and record metadata.

The *pyPhasesML* plugin provides an interface to handle specific preprocessing steps, data manipulation and methods to train machine learning models. It handles the machine learning pipeline: extracting data, efficiently loading data, manipulating data and training models. At the moment, *pyTorch* and *tensorflow* models are supported.

*SleePyPhases*, *pyPhases*, *SleepHarmonizer*, *pyPhasesML*, all specific record loaders, packages and the code to reproduce the experiments are made publicly available under the MIT License on the *Python Package Index* and *GitLab*, see https://gitlab.com/sleep-is-all-you-need/sleepyphases, https://gitlab.com/sleep-is-all-you-need/pyphases and https://gitlab.com/sleep-is-all-you-need/reproduce/. This public availability ensures the framework follows FAIR principles.

#### 2.3.3 Data Acquisition

Data acquisition is provided by the pyPhasesRecordloader module. Each specific

*RecordLoader* loads PSG data using three methods:

- getSignals returns a *RecordSignal* which is a composition of signals including information such as the sampling rate and the signal itself as NumPy array
- getEventList returns a list of events and annotations with start times and durations
- getMetaData returns a key-value dictionary of record- and channel-specific metadata

Several predefined recordloaders are available to load data from different datasets including Sleep Heart Health Study (SHHS) (Quan et al. 1997), Multi-Ethnic Study of Atherosclerosis (MESA) (Chen et al. 2015), MrOS Sleep Study (MrOS) (Blackwell et al. 2011), George B. Moody PhysioNet Challenges 2018 (PhysioNet) (Ghassemi et al. 2018), SleepEDF Database Expanded (SleepEDF) (Kemp et al. 2000), Human Sleep Project (HSP) (Westover et al. 2023) and two different vendors including Philips Alice® and Somnomedics Domino®. A loader can be activated by setting the configuration value useLoader to the name of the loader (e.g. useLoader: shhs). For a predefined dataset recordloader, only the path is needed (e.g shhs-path: /dataset/shhs).

To include custom setups or new datasets, the file structure needs to be defined to extract the recordID patterns. Additionally, the channel configuration is required in the configuration. All loader configurations are stored in the main parent loader configuration. A configuration to handle the shhs dataset which is included in the pyPhasesRecordloaderSHHS module is shown in Listing 1.

Listing 1: Example Configuration to handle SHHS dataset

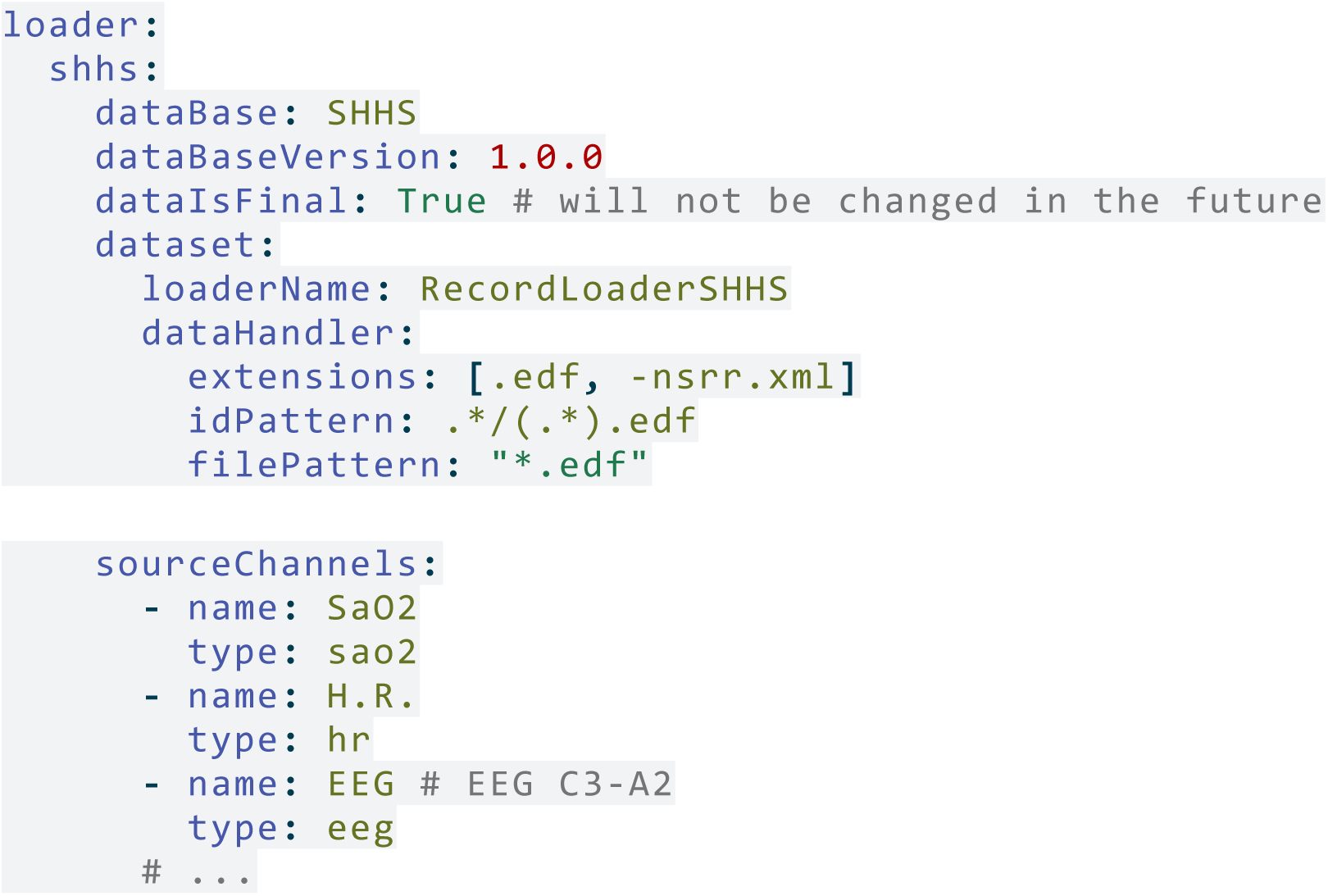

The data can be loaded at any phase using the RecordLoader Singleton and the loadRecord method as shown in Listing 2. This method returns a *RecordSignal*, which contains metadata and all signals.

Listing 2: Loading an EEG from a specific SHHS-record using the RecordLoader Singleton

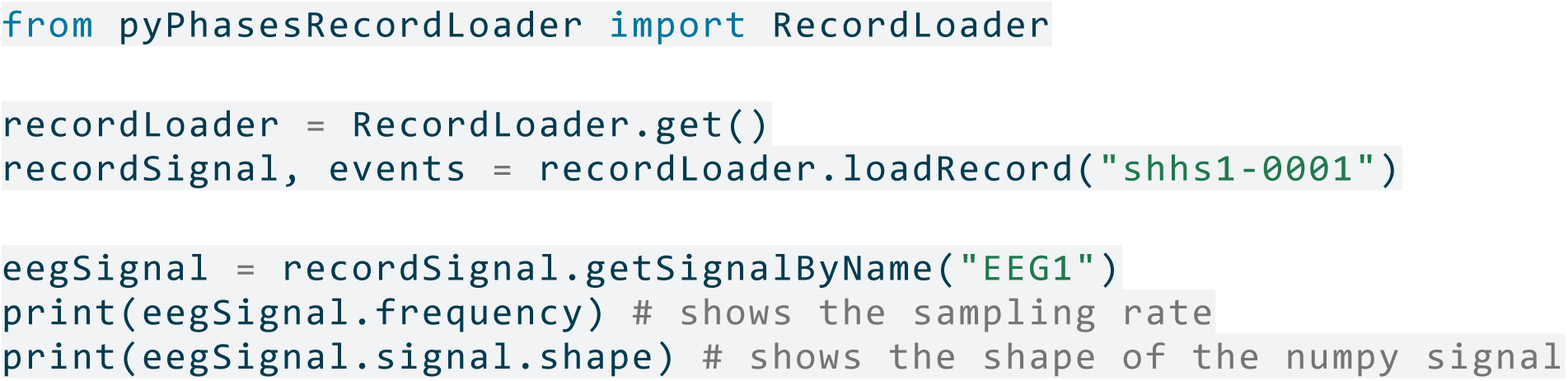

#### 2.3.4 Filtering and splitting the data

The data can be filtered using the dataVersion configuration. This allows records to be specified by ID and filtered by record or channel metadata. Filtering is done using the metadata which is a pandas dataframe with all recordIds and given metadata. A filterQuery filters the records by the given query. An example to filter all SHHS records from the first part of the study with an Apnea-Hypopnea-Index greater than 15 is shown in Listing 3. Additional splits can be defined using the split configuration, to specify training, validation and test splits. These splits can be explicit or be relative by defining a fold or with values from zero to one for validationSplit and testSplit.

Listing 3: Filtering SHHS1 records with an apnea-hypopnea index (AHI) greater than 15. The records are shuffled using the seed 2025 and split into a holdout testset and four training-validation folds.

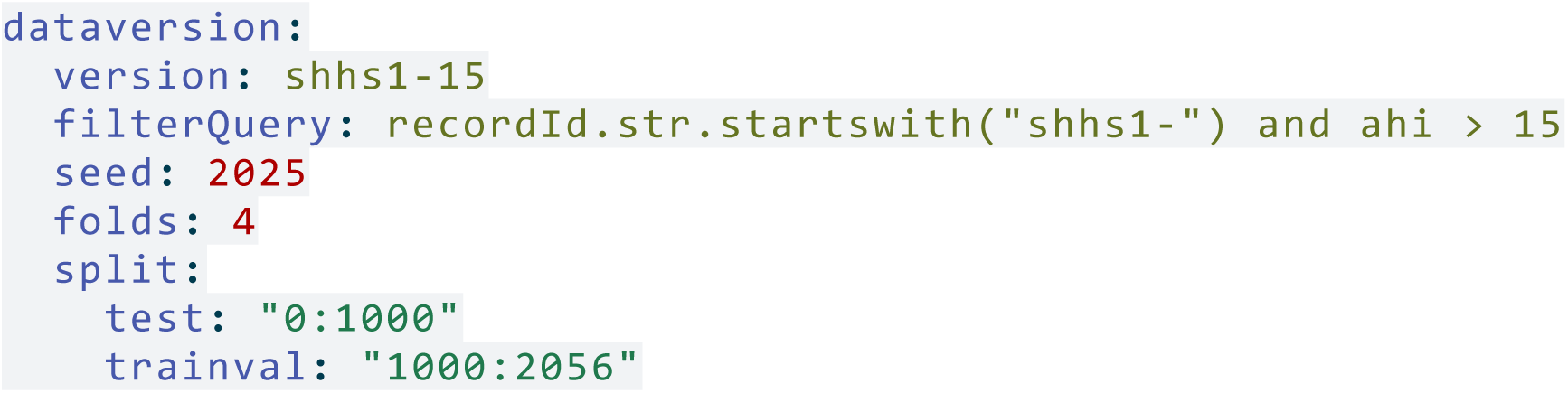

A list of records can be loaded within a *Phase* using python self.getData(’allRecordIds’, list) and separate splits by using the dataversionmanager as shown in Listing 4.

Listing 4: Loading all test record IDs using the DataVersionManager

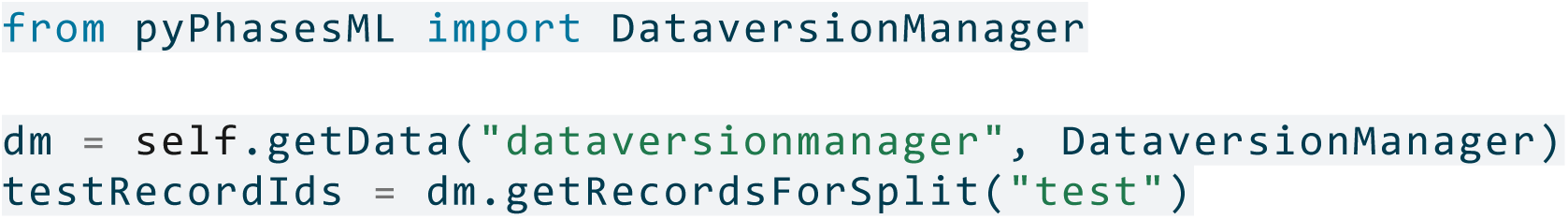

#### 2.3.5 Data Preprocessing

The data preprocessing is done using the preprocessing and labelChannels configuration. This allows to define which label channels and target channels should be used. The preprocessing includes a target frequency for signals and labels, manipulation of the whole record and events (*PreManipulation* using the config manipulationSteps) and preprocessing steps per type (stepsPerType). Additionally, the configuration featureChannels allows to add additional channels calculated from the *RecordSignal*, that can be used as target channels. The preprocessed data is stored in a *NumPy* memory map, which allows efficient data loading. An example on loading a single record can be found in Listing 7. How to resample and standardize a photoplethysmography (PPG) signal is shown in Listing 5. As there are multiple ways to preprocess the data, the toolbox provides a flexible configuration system that allows to define custom manipulation and preprocessing steps as shown in Listing 6.

Listing 5: Preprocessing configuration to resample PPG signals to 64Hz and standardize them.

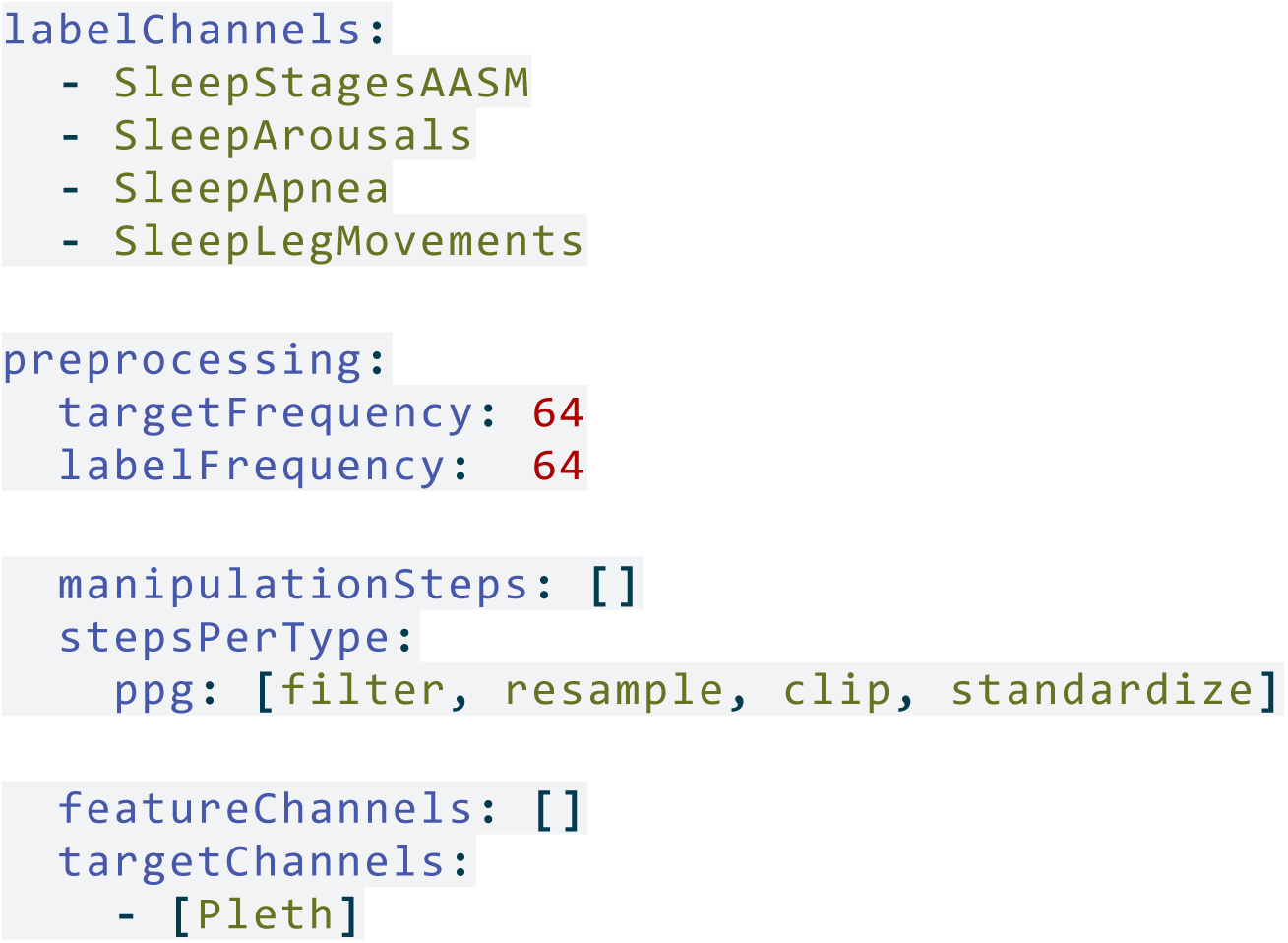

Listing 6: Custom preprocessing step to standardize a signal

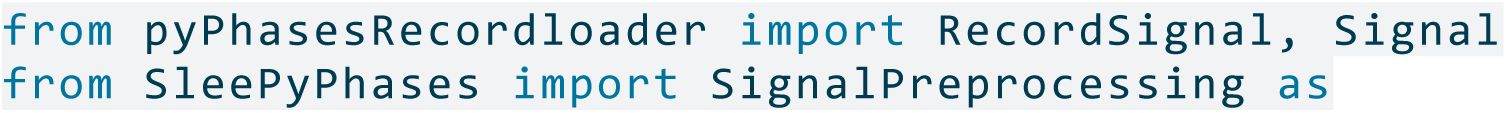
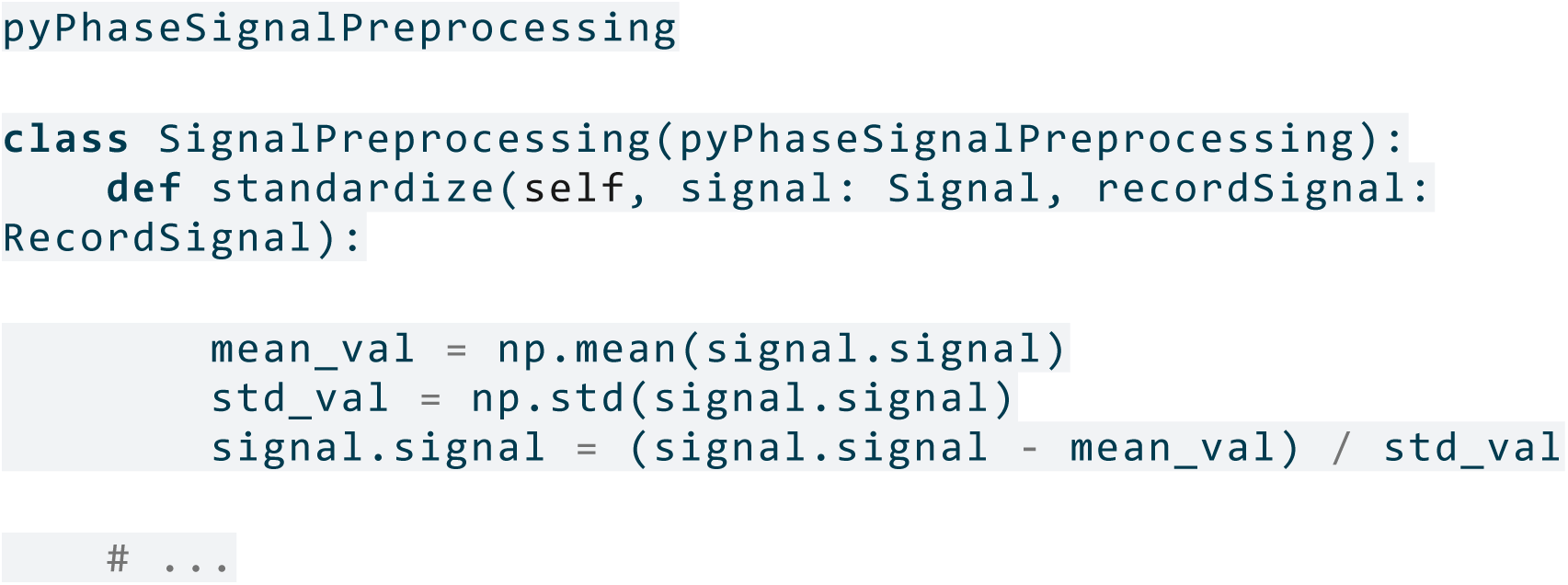

Listing 7: Loading the first record of an extracted dataset

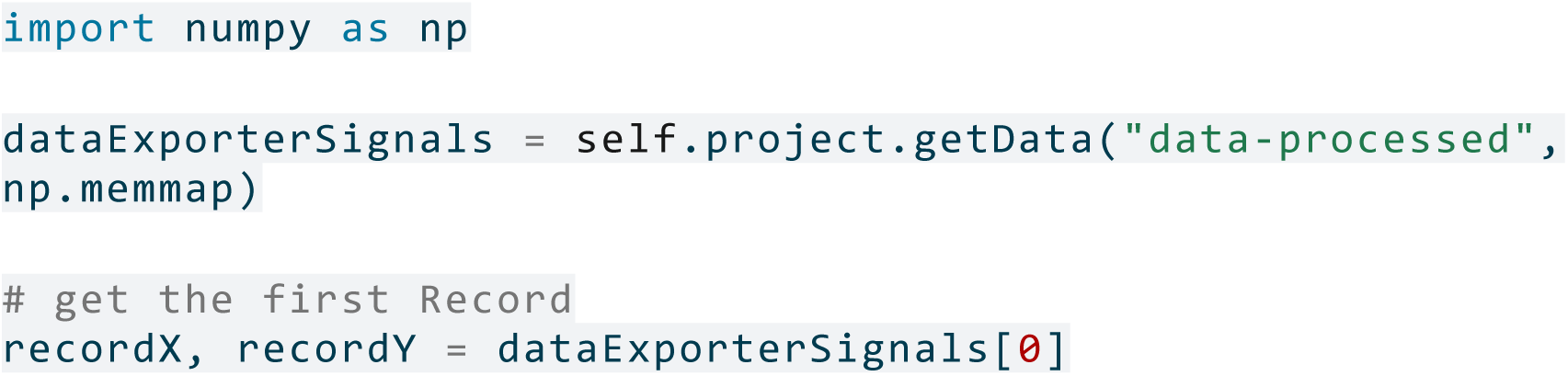

#### 2.3.6 Data Manipulation

Data manipulation is the step from loading the optimized data (preprocessing) and augmenting or manipulating it before being processed by the algorithm. The manipulation phase transforms the preprocessed data into machine learning model input. A typical example of manipulation is data augmentation to generate better training data, but also tensor transformation to generate the suitable input shape. The manipulation is done using several configuration points:

- segmentManipulation: executed on a single segment of a signal or complete recording (depending on the recordWise configuration)
- batchManipulation: executed on a batch of segments of a signal, before it is passed to the machine learning algorithm.
- manipulationAfterPredict: executed on the predictions of the machine learning algorithm, before they are returned to the user.

Listing 8: Example configuration for manipulation steps on segments, batches and predictions.

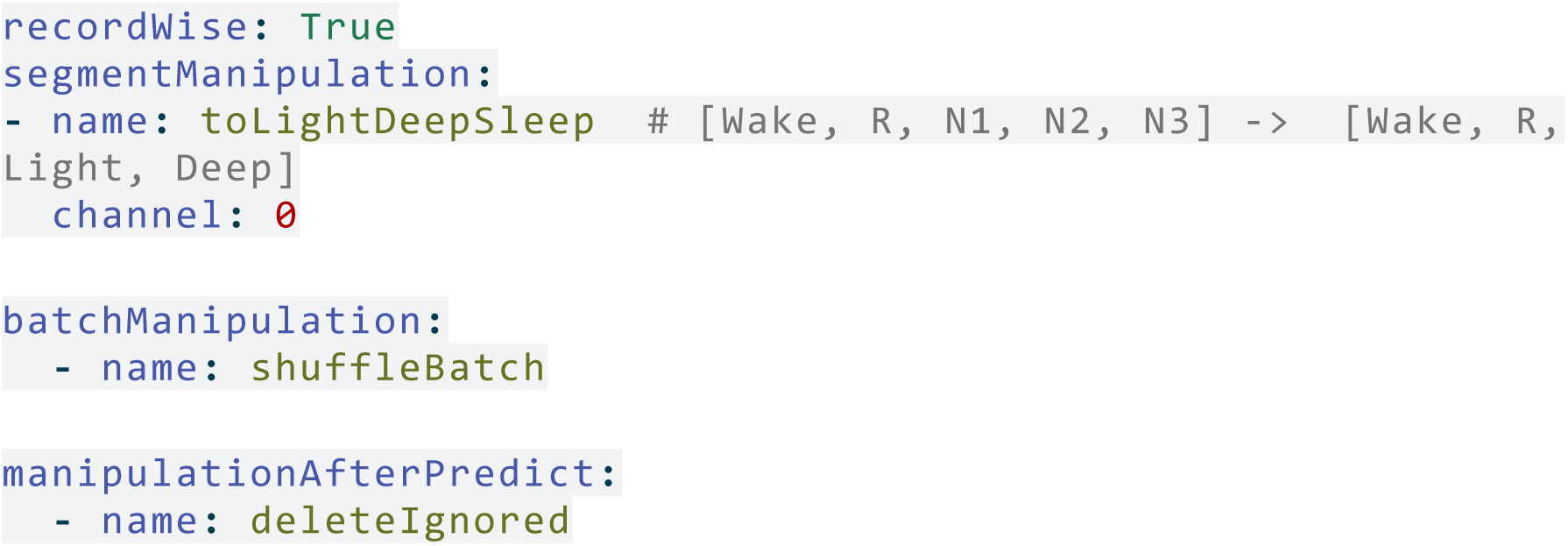

Similar to the signal preprocessing, the manipulation configuration allows to define custom manipulation steps (*DataManipulation*) as shown in Listing 8, and their implementation in Listing 9.

Listing 9: Custom manipulation step to convert N1, N2 and N3 to light and deep Sleep, shuffle the batch, and ignore all undefined annotations after prediction.

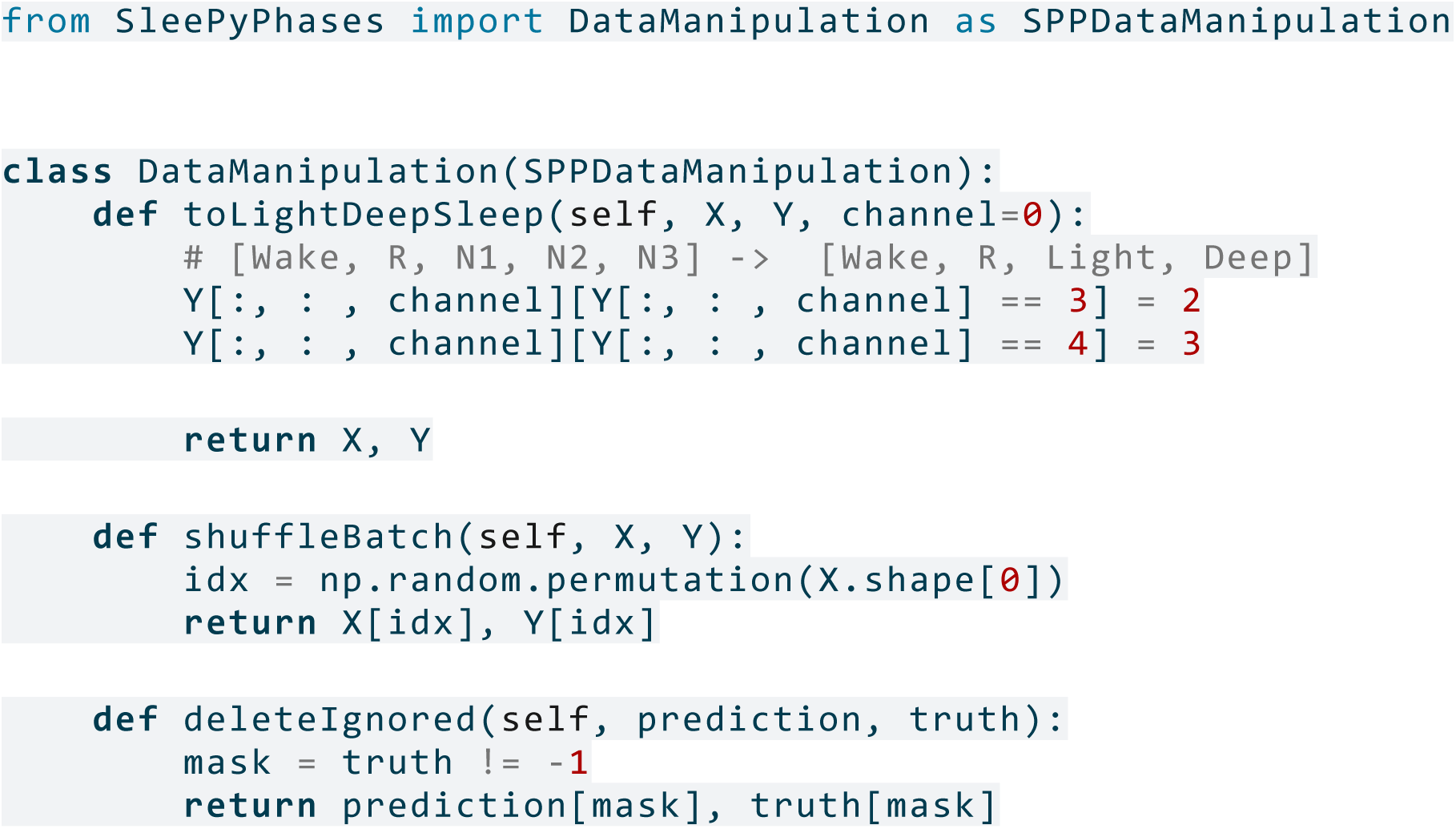

#### 2.3.7 Evaluation and Reporting

The evaluation is divided into two parts: a segment-wise evaluation and an event-wise evaluation. The segment-wise evaluation scores the evaluation metrics on the test set segments defined by the preprocessing.targetFrequency configuration. The event-wise evaluation combines consecutive segments with the same label into a single event. Depending on an overlap, the confusion matrix can then be calculated. For event evaluation, threshold optimization can be performed to determine the optimal threshold for the given evaluation metric on the validation set.

The event evaluation is stored with performance metrics and calculated PSG parameters for each recording in a comma separated values (CSV) file, and a summary of the evaluation is stored in a results.yml file. The Listing 10 shows a configuration that uses different metrics for sleep scoring and calculates PSG parameters on a record-by-record basis.

Listing 10: Example configuration for using different evaluation metrics on different scoring components and for calculating clinical metrics.

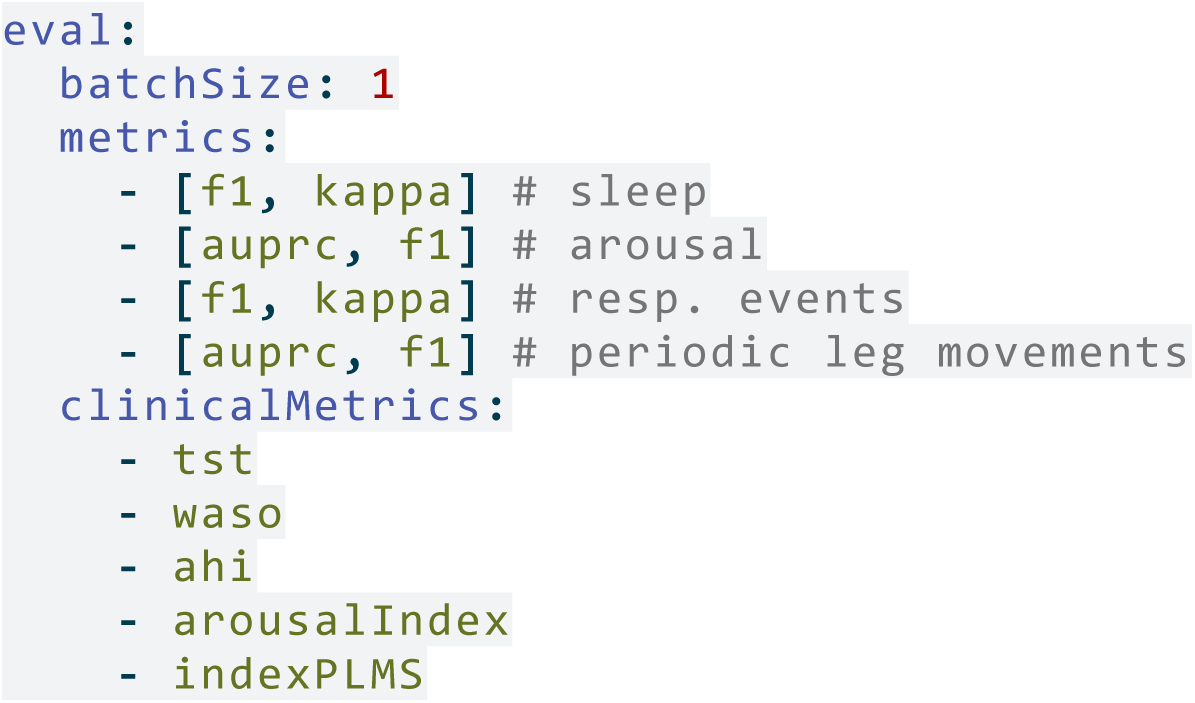

## 3. Results

### 3.1 Framework Validation Through Study Reproduction

All five selected studies were successfully reproduced using the harmonized data access workflow, demonstrating that standardized repository access can replace custom data handling implementations. Each reproduction validates specific aspects of the framework’s data harmonization capabilities while confirming that the framework maintains data fidelity across diverse access patterns.

The implementation of the DRCNN by (Pourbabaee et al. 2019) with *SleePyPhases* (SPP-DRCNN) validates the framework’s ability to access challenge-based datasets with specific formatting requirements. Accessing PhysioNet 2018 data through *pyPhasesRecordloaderPhysionet* required harmonizing 12 different PSG channel types and complex annotation schemas. The reproduction demonstrates that standardized data loading produces identical results to custom implementations, validating that the harmonization layer preserves data integrity for multi-channel recordings with diverse sampling rates and challenge-specific annotation formats.

The SleepTransformer reproduction (Phan et al. 2022) (SPP-SleepTransformer) validates the framework’s multi-repository access capabilities. Successfully accessing both SHHS and SleepEDF through their respective loaders (*pyPhasesRecordloaderSHHS*, *pyPhasesRecordloaderSleepEDF*) demonstrates that the framework provides consistent data access across repositories with different organizational structures and metadata formats. The near-identical reproduction results confirm that standardized access produces functionally equivalent data regardless of the underlying repository structure, enabling seamless multi-dataset studies without custom data handling code.

The SleepPPG reproduction (Kotzen et al. 2022) (SPP-SleepPPG-Net) validates the framework’s capability for signal-specific access and three-way repository harmonization. Integrating data from MESA, SHHS, and CFS through unified interfaces (*pyPhasesRecordloaderMESA*, *pyPhasesRecordloaderSHHS*, *pyPhasesRecordloaderCFS*) demonstrates that the framework enables direct comparison and inter-dataset validation studies. The reproduction confirms that PPG-specific preprocessing pipelines can be consistently applied across repositories, validating the framework’s ability to handle specialized signal types and cross-dataset research designs.

The MSED reproduction (Zahid et al. 2023) (SPP-MSED) validates the framework’s extensibility for complex event-based data access patterns. Accessing MrOS data with custom segmentation strategies demonstrates that the framework accommodates non-standard data access requirements while maintaining standardization. The successful reproduction confirms that even complex, study-specific data extraction patterns can be implemented within the standardized workflow, validating its flexibility for diverse research methodologies.

The NeuroNet reproduction (Lee et al. 2024) (SPP-NeuroNet) validates the framework’s support for multi-stage, multi-dataset access patterns required by self-supervised learning approaches. Accessing SleepEDFX and SHHS data through separate training phases demonstrates the framework’s ability to manage complex data dependencies and cross-dataset transfer learning scenarios. While this reproduction encountered challenges due to incomplete augmentation specifications in the original publication, the partial success still validates that the framework correctly accesses and harmonizes data across multiple repositories for advanced training strategies.

### 3.2 Quantitative Validation of Data Access Fidelity

The reproduction results provide quantitative validation that the harmonized data access workflow produces functionally equivalent data to original implementations. Near-identical performance metrics confirm that standardized repository access preserves data integrity and does not introduce artifacts or systematic errors.

The successful reproduction of all five studies using the *SleePyPhases* framework validates its capability to provide reliable, standardized access to diverse sleep data repositories. The reproduction results, summarized in Table 2, demonstrate data access fidelity across multiple repositories and methodological approaches.

**Table 2:**
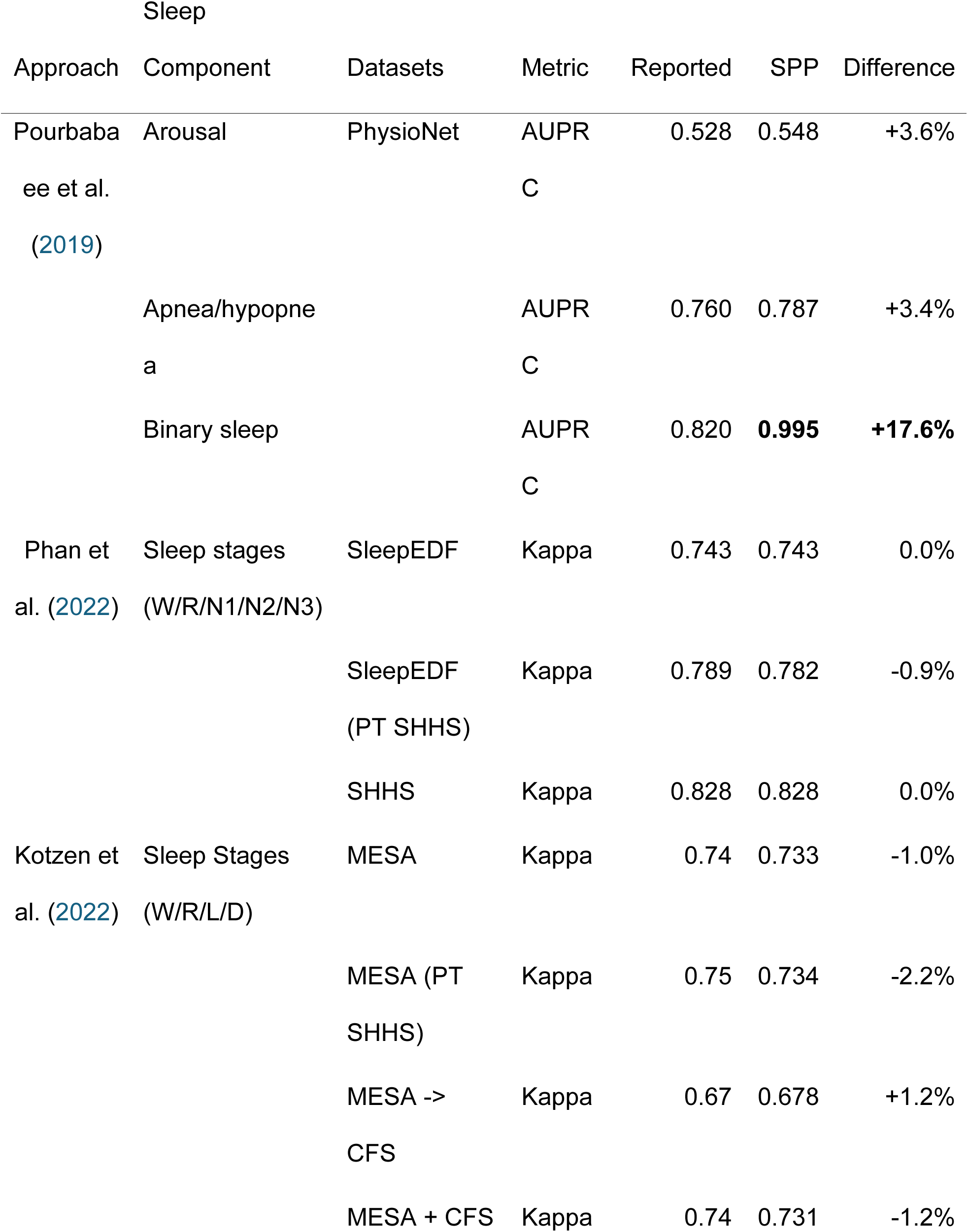
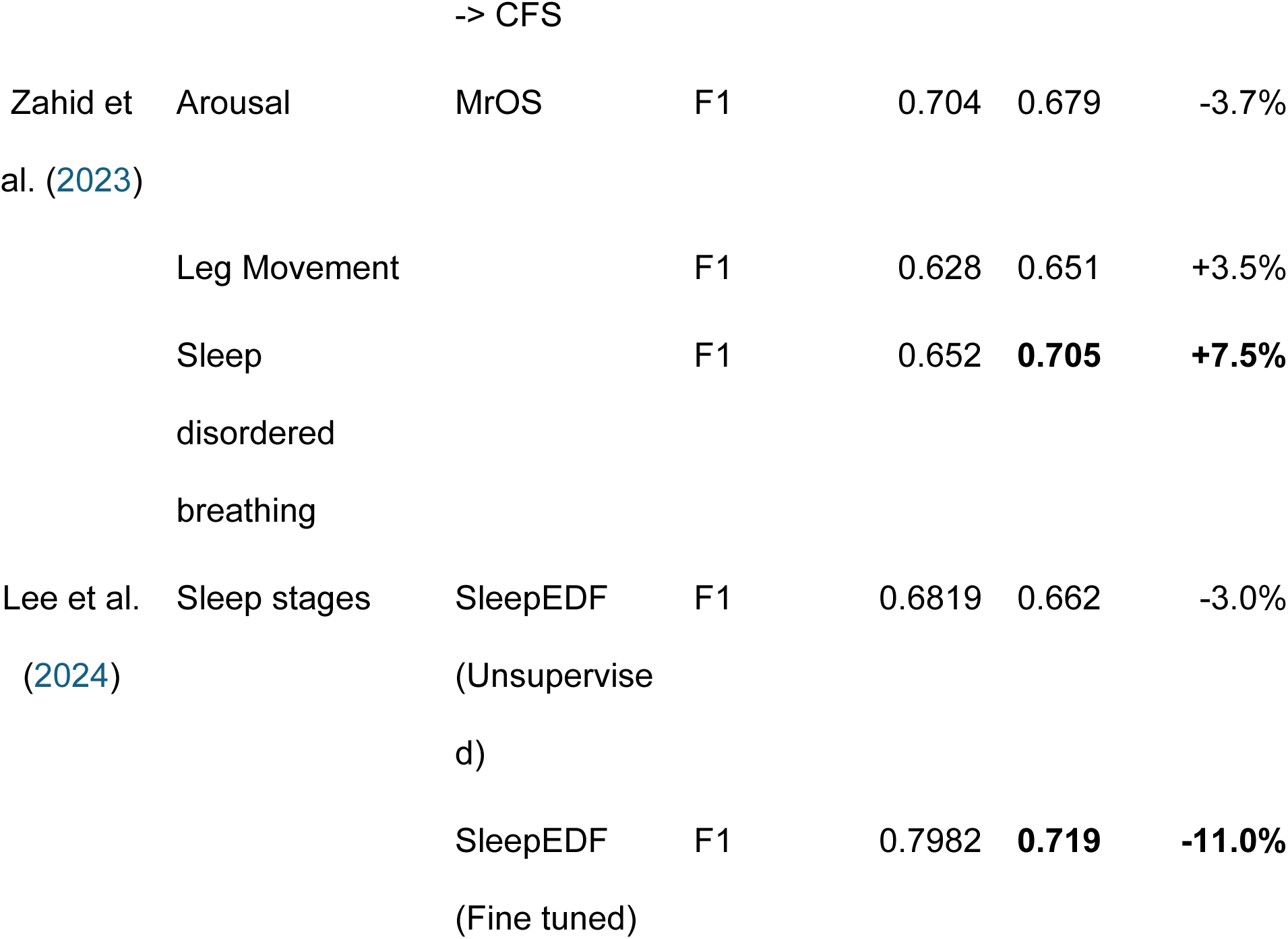
Results of reproducing the different approaches from the literature versus implementing them with the SleePyPhases (SPP) framework. Results with a relative difference of more than 5% are in bold. CFS: Cleveland Family Study Visit-5 v1, SHHS: Sleep Heart Health Study, MESA: Multi-Ethnic Study of Atherosclerosis, MrOS: MrOS

Four of the five reproductions produced results that closely matched (less than 5% deviation) those in the original publications. This confirms that: (1) the harmonized data access workflow correctly loads data from repositories, (2) the original methods function correctly with standardized data access, (3) the framework-based implementations now provide open-source alternatives to previously closed methods, and (4) standardized access enables reproducibility that was previously impossible. Performance differences relative to original results are reported as percentage changes, with positive values indicating higher performance and negative values indicating lower performance. Detailed comparisons are provided in Table 2.

Replicating Pourbabaee et al. (2019) demonstrates successful harmonized access to PhysioNet 2018 data, achieving similar results for arousal detection (+3.6%) and apnea prediction (+3.4%), confirming that standardized data loading preserves multi-task annotation fidelity. The notably improved binary sleep classification performance (+17.6%) suggests the framework may actually improve data quality through consistent preprocessing, though the source of improvement requires further investigation. Reproducing Phan et al. (2022) validates cross-repository access fidelity with nearly identical results across all conditions: *SleepEDF* (±0.0%), *SleepEDF* pretrained on *SHHS* data (-0.9%), and *SHHS* training (±0.0%). This confirms that the framework provides equivalent data access across repositories with different structures. The Kotzen et al. (2022) reproduction validates three-way repository harmonization with close results (-1.0% for *MESA* and -2.2% for *MESA* pretrained on *SHHS*), confirming that standardized access enables seamless cross-dataset validation studies. Dataset cross-validation also yielded similar results (+1.2% for CFS trained on MESA), validating the framework’s support for inter-dataset transfer scenarios. The Zahid et al. (2023) reproduction validates custom event-based access patterns with similar results for arousal detection (-3.7%) and leg movement detection (+3.5%). The sleep disordered breathing detection improvement (+7.5%) may reflect enhanced event alignment through standardized access. The Lee et al. (2024) reproduction showed similar self-supervised learning performance (-3.0%), though the fine-tuned model (-11.0%) revealed that incomplete parameter documentation in the original publication limits full reproducibility—highlighting how the framework enables transparency by exposing such gaps through open implementation attempts.

## 4. Discussion

The SleePyPhases workflow framework has been successfully validated across multiple databases and methods, supporting a variety of model architectures and preprocessing approaches within a unified, configuration-driven environment. Reproducing five different sleep analysis algorithms highlights the framework’s capabilities and the challenges of creating reproducible machine learning pipelines for sleep research. The capabilities are complemented by our previous research, where (Ehrlich et al. 2024) used the Philips Alice® 6 as custom vendor format and (Goldammer et al. 2022) used the TensorFlow machine learning framework.

All selected validation studies were successfully reproduced using standardized data access, demonstrating the framework’s versatility in handling diverse repository structures, annotation schemas, and data organization patterns. The framework enables researchers to focus on scientific questions and methodological development rather than low-level data infrastructure concerns.

The reproduction studies systematically validated the six core framework requirements. **Data Harmonization** was demonstrated through successful standardized access to multiple repositories with different organizational structures and vendor formats. This remains an ongoing community effort as new repositories and vendors emerge. While we did not focus on the internal *DataSchema* (Cheng et al. 2024) in this research, we provide a function to export the PSG using standardized codings for annotations and signal representations. **Configurable Preprocessing** was validated through diverse preprocessing pipelines applied consistently across repositories, from sliding normalization windows (DRCNN) to spectrogram extraction (SleepTransformer). Addionally, the framework’s memory-mapped storage system provides efficient access while maintaining configuration-based provenance. A configuration-based method was used to define implementations, achieving **Flexible Data Manipulation** ranging from default event windows (MSED) to complex augmentation strategies (NeuroNet). Reproduction studies have demonstrated the successful application of multiple analysis frameworks (e.g. PyTorch and TensorFlow) and training strategies (e.g. supervised and self-supervised) within the framework. Therefore, the **Integrated Training Pipeline** requirement has been met. The pyPhases component ensured the **Configuration-Data Synchronization**, through explicit configuration-based provenance that connects data access patterns to results and ensuring experiment reproducibility. Finally, the **Modular Architecture** was validated through the independent development of repository-specific loaders that can be shared and by different research groups without any need to modify the core framework code. Therefore, the framework meets all the requirements identified in the literature (Cheng et al. 2024) and based on our expertise.

The framework’s design inherently supports the FAIR principles (Wilkinson et al. 2016) for research data and software. The framework is **Findable** through public repositories (GitLab, Python Package Index). It is **Accessible** via open-source MIT licensing, enabling unrestricted use and modification. **Interoperability** is achieved through data harmonization that support multiple signal formats (open formats and proprietary vendor formats), channel naming conventions, and harmonized annotation semantics across diverse datasets. Finally, the framework ensures **Reusability** through its modular plugin architecture, versioned releases, extensive configuration options, and reproduction of existing methodologies. Adherence to the FAIR principles is a step towards more transparent and collaborative sleep research.

Our validation efforts yielded excellent results, confirming data access fidelity. Four studies (DRCNN (Pourbabaee et al. 2019), SleepTransformer (Phan et al. 2022), SleepPPG-Net (Kotzen et al. 2022), and MSED (Zahid et al. 2023)) achieved near-identical results using standardized data access compared to original custom implementations, validating the framework’s data integrity. Importantly, these reproductions now provide open-source alternatives to methods that were previously unavailable, advancing research transparency.

The DRCNN reproduction produced notably improved binary sleep classification results (AUPRC: 0.820→0.995). Since standardized data access is the primary change from the original implementation, this suggests the framework’s consistent preprocessing may enhance data quality, though the specific mechanisms require further investigation. We examined early validation results and found no anomalies suggesting the improvement is spurious.

The MSED study comprises three tasks: sleep arousal, sleep-disordered breathing (SDB), and leg movements. The reproduced SDB task showed an increase of around 8%, while performance on the arousal task was slightly lower (-3.7%). During training, the optimal model weights were selected based on overall loss (the sum of losses for all tasks). Training logs indicate that arousal task performance peaked earlier in the validation set, potentially explaining the discrepancy between original reported values and our results.

Despite the training process and model architecture being open source, the replication of the NeuroNet study illustrates how standardized data access workflows can reveal documentation gaps in published research. The self-supervised learning (SSL) model produced slightly lower performance (-3%) than reported. As no detailed parameters were provided for data augmentation—a crucial component for SSL—we suspect incomplete method documentation rather than framework limitations. The standardized data loading provided by SleePyPhases ensures the discrepancy stems from undocumented hyperparameters, not data access issues. This transparency benefit demonstrates how workflow standardization can improve research reproducibility by isolating methodological ambiguities.

The NeuroNet reproduction highlights the value of open-source workflow implementations for scientific transparency. While the original study provided model architecture code, the lack of complete training configuration parameters prevented exact reproduction. By providing this open implementation through SleePyPhases, future researchers gain access to both the data handling and a documented reference implementation, addressing gaps in the original publication. This represents a key contribution: converting proprietary or incompletely documented research into fully transparent, reproducible implementations.

Several design considerations emerged from the reproduction studies. The SleepTransformer reproduction required adapting intra-epoch validation to full epoch-based validation due to framework architecture, though this did not affect final results. The framework’s TensorFlow 2.0+ requirements necessitated model conversions for some legacy implementations, highlighting version compatibility considerations when providing long-term data access infrastructure. These adaptations demonstrate the framework’s flexibility while identifying areas where enhanced abstraction could improve methodological fidelity across diverse study designs.

The workflow framework provides efficient infrastructure for multi-dataset sleep research. The combination of optimized preprocessing storage and flexible data manipulation enables rapid iteration during method development. The configuration-based approach allows researchers to modify experimental parameters without code changes, facilitating systematic exploration of data access patterns and preprocessing strategies.

The framework’s ability to handle diverse datasets and preprocessing requirements through configuration alone significantly reduces the technical overhead typically associated with multi-dataset studies. This standardization enables researchers to focus on scientific questions rather than data infrastructure concerns, lowering barriers to entry for sleep research and enabling studies that would be impractical with custom data handling approaches.

Although the framework is primarily focused on sleep research, the principles are applicable to other physiological signal analysis domains. For example, similar harmonized data access patterns could support ECG analysis for arrhythmia detection or EEG analysis for seizure detection when appropriate dataset plugins are provided.

## 5. Conclusion

The *SleePyPhases* framework addresses critical challenges in sleep research by providing unified access to different sleep data repositories and custom PSG setups. It also provides a configuration-driven platform for data analysis and the development of machine learning pipelines. Our work demonstrates that standardization and harmonization of sleep data processing can be achieved without sacrificing methodological flexibility or algorithmic innovation. The integrated data harmonization proves particularly valuable for multi-dataset studies. Future development should focus on enhancing accessibility through improved documentation and tutorials, and expanding support for more intuitive extension points.

The successful reproduction of four published studies with near-identical results validates the framework’s data integrity. Importantly, these reproductions now provide open-source implementations of methods that were previously unavailable, advancing research transparency and enabling community validation. The framework’s memory-mapped storage system and efficient preprocessing pipeline significantly reduce technical overhead for multi-dataset studies while maintaining methodological flexibility.

*SleePyPhases* represents a step toward FAIR-compliant sleep research infrastructure. By providing standardized access to multiple data repositories, the framework reduces barriers to multi-dataset research and enables studies that would be impractical with custom data handling approaches. The open-source availability under the MIT license, combined with the modular plugin architecture, ensures that the community can extend support to new repositories and contribute improvements. While this implementation focuses on sleep research, the framework principles can be adapted to other physiological signal analysis domains requiring multi-repository access.

## 6. Code and Data Availability

All source code is made available under the MIT License on GitLab:

- **SleePyPhases Framework**: https://gitlab.com/sleep-is-all-you-need/SleePyPhases - Main framework implementation
- **Sleep Harmonizer**: https://gitlab.com/sleep-is-all-you-need/sleep-harmonizer - PSG data harmonization plugin
- **Reproduction Studies**: https://gitlab.com/sleep-is-all-you-need/reproduce - All reproduction experiments as sub-projects
- **pyPhases Core**: https://gitlab.com/sleep-is-all-you-need/pyphases - Core pyPhases framework and all related projects including pyPhasesRecordLoader and dataset-specific record loaders (SHHS, MESA, MrOS, etc.)

The SHHS, MESA and MrOS datasets for validating the framework are available from the National Sleep Research Resource website (https://sleepdata.org/datasets), while the PhysioNet 2018 and Sleep-EDF datasets are available from Physionet (https://physionet.org/).

## 7. Funding

This research was funded by the the Federal Ministry of Research, Technology and Space under the funding code 01ZZ2324F.

## Data Availability

All source code is made available under the MIT License on GitLab: - **SleePyPhases Framework**: (https://gitlab.com/sleep-is-all-you-need/SleePyPhases) - Main framework implementation - **Sleep Harmonizer**: (https://gitlab.com/sleep-is-all-you-need/sleep-harmonizer) - PSG data harmonization plugin - **Reproduction Studies**: (https://gitlab.com/sleep-is-all-you-need/reproduce) - All reproduction experiments as sub-projects - **pyPhases Core**: (https://gitlab.com/sleep-is-all-you-need/pyphases) - Core pyPhases framework and all related projects including pyPhasesRecordLoader and dataset-specific record loaders (SHHS, MESA, MrOS, etc.) The SHHS, MESA and MrOS datasets for validating the framework are available from the National Sleep Research Resource website (https://sleepdata.org/datasets), while the PhysioNet 2018 and Sleep-EDF datasets are available from Physionet (https://physionet.org/).

https://sleepdata.org/datasets

https://gitlab.com/sleep-is-all-you-need/SleePyPhases

https://gitlab.com/sleep-is-all-you-need/sleep-harmonizer

https://gitlab.com/sleep-is-all-you-need/reproduce

https://gitlab.com/sleep-is-all-you-need/pyphases

## Acknowledgements

The authors gratefully acknowledge the computing time made available to them on the high-performance computer at the NHR Center of TU Dresden. This center is jointly supported by the Federal Ministry of Education and Research and the state governments participating in the NHR (www.nhr-verein.de/unsere-partner).

## 8. Apendix

Listing 11: Example Configuration for a basic pyPhases project

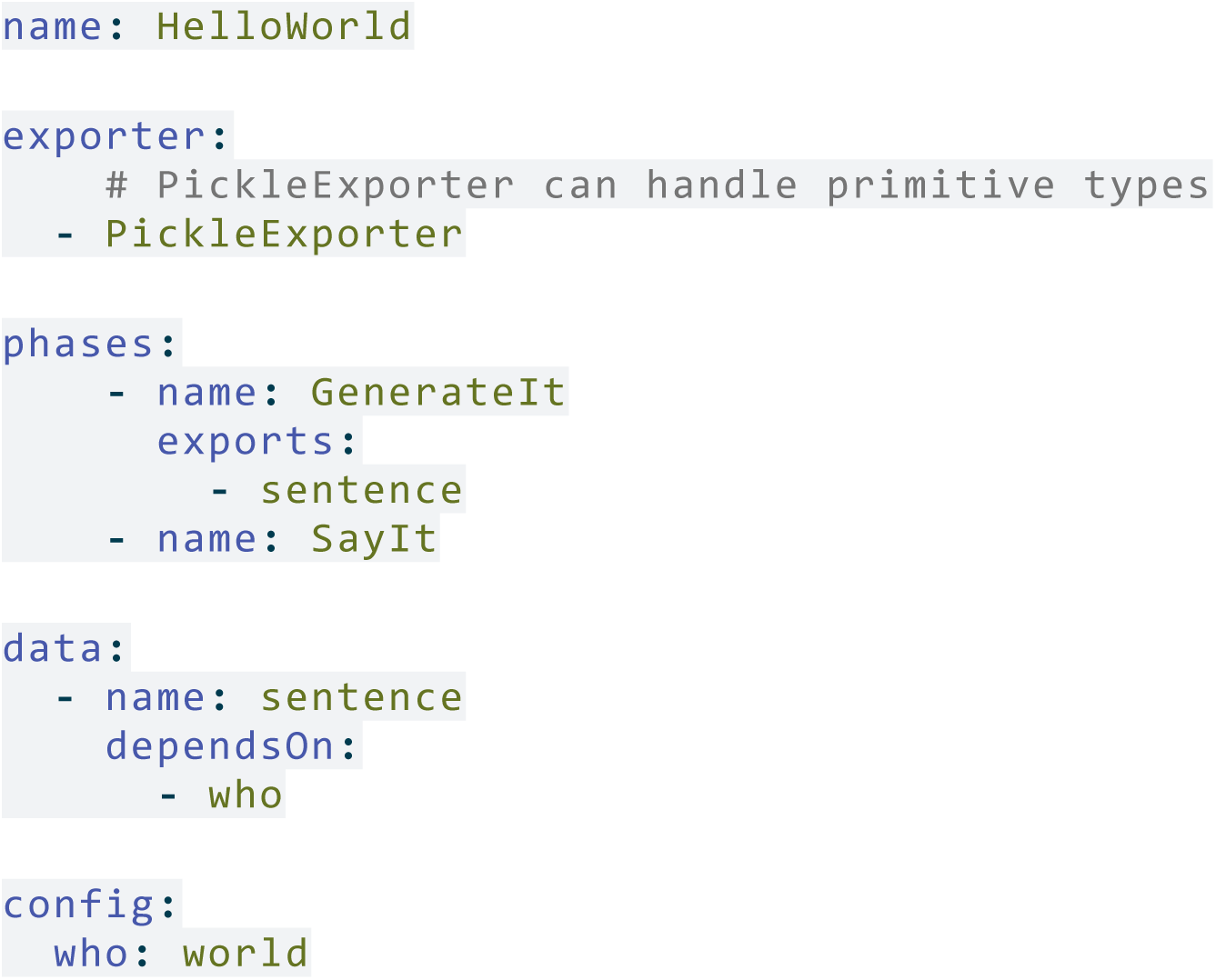

Listing 12: Example phase to generate data for the basic pyPhases project

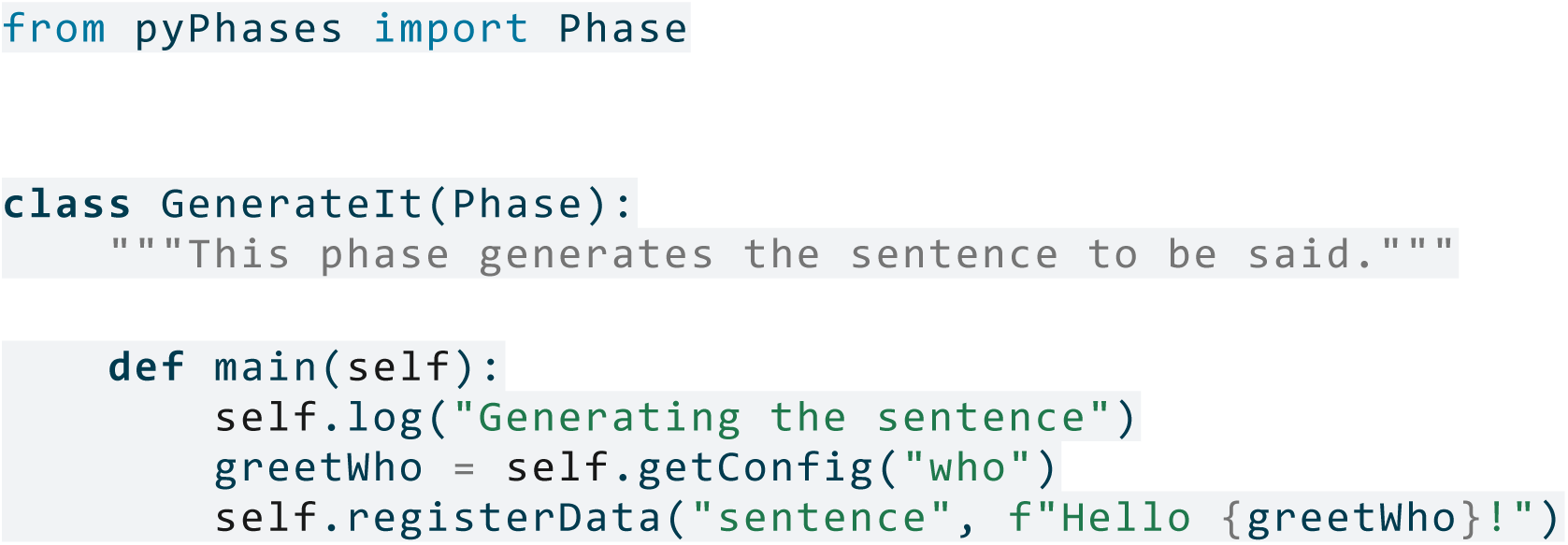

Listing 13: Example phase to load data for the basic pyPhases project

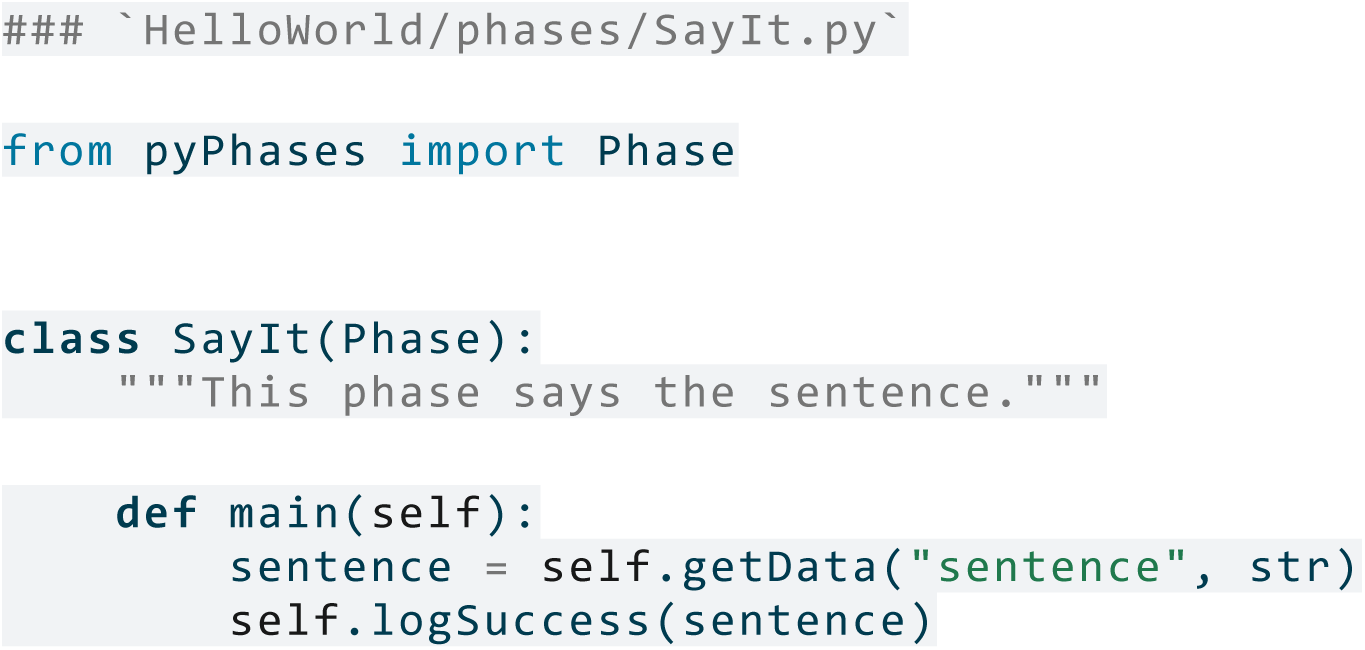

## Notes

### Competing Interest Statement

The authors have declared no competing interest.

### Funding Statement

Yes

### Author Declarations

This study did not require Institutional Review Board approval because it involved secondary analysis of publicly available, de-identified data.

